# “Effect of the intervention with omega-3 fatty acids on nutritional and clinical aspects in patients with breast cancer receiving medical treatment”

**DOI:** 10.1101/2021.02.08.21251389

**Authors:** Palma-Gutierrez Edgardo, Espinoza-Rado Erika, Zafra-Tanaka Jessica Hanae

## Abstract

**Background:** It is known that cancer can cause loss of body weight and muscle protein wasting, which leads to a state of malnutrition, which in turn worsens the prognosis and health of the cancer patient. It has been suggested that the promoting mechanism of this state is systemic inflammation, for which reason several clinical trials have used omega-3 fatty acids, especially eicosapentaenoic acid (EPA) and docosahexaenoic acid (DHA), as adjuvants to antineoplastic treatment, mainly due to its anti-inflammatory effects. However, few systematic reviews and meta-analyzes have analyzed the effects of omega-3s in patients with breast cancer.

**Objective:** The aim of this study is to assess the effect of the supplementation with omega-3 fatty acids on nutritional and clinical outcomes in patients with breast cancer receiving medical treatment.

**Methods:** A systematic review will be conducted, starting with a search in PubMed, CENTRAL and EMBASE using search terms related to omega-3 fatty acids and breast cancer. We will include only randomized controlled trials that assess the effects of omega-3 in patients with breast cancer receiving medical treatment.. Data will be extracted in a spread sheet. Study selection and data extraction will be conducted by two reviewers independently and the Cochrane Risk of Bias Tool for RCT will be used for assessment of risk of bias. Discrepancies will be reviewed with a third reviewer.

**Conclusion:** This systematic review aims to provide an analysis on the outcomes of the usage of the intervention with omega-3 fatty acids on nutritional and clinical aspects in patients with breast cancer receiving medical treatment.

## Introduction

Breast cancer has one of the highest burdens of disease among cancer. In 2018, it affected around 2 261 419 women worldwide and caused the death of 684 996 patients.^1^. It is known that cancer can lead to severe malnutrition; possibly, due to the systemic inflammation that it promotes that induces loss of weight and muscle mass, which in turn worsens the prognosis of the patient^2^. Malnutrition can be very frequent and has a high impact on these people, since it has been estimated that 10 to 20% of deaths from cancer may be due to malnutrition rather than to the neoplasia itself^2^. In other studies, worse response rates to chemotherapy drugs were observed in patients who lost weight, including women with breast cancer^3^.

It is known that omega-3 fatty acids could help fight malnutrition associated with cancer, due to their anti-inflammatory effects that reduce the breakdown and wasting of muscle mass and improve other nutritional aspects^4,5^. The types of omega-3 most studied in antineoplastic therapy have been eicosapentaenoic acid (EPA) and docosahexaenoic acid (DHA). Some clinical trials have reported that omega-3 supplementation helps modulate the immune response^6^, and improved the survival of breast cancer patients receiving antineoplastic treatment^7^. With regards to nutritional outcomes, no significant differences were found on weight or body composition between those who received the supplementation and those who did not^8^.

There are physiological mechanisms by which omega-3 could be part of the treatment of breast cancer. However, the effects of this intervention on nutritional and clinical outcomes remains unclear. Moreover, there is a need to further explore how the dose, the time of intervention and the characteristics of the population can modify the effects of omega-3. The objective of this systematic review and meta-analysis will be to analyze the effect of the supplementation with omega-3 fatty acids, in their form of EPA and DHA, on nutritional and clinical outcomes in patients with breast cancer receiving medical treatment.

## Methods

### Study design

This systematic review will be conducted according to the preferred reporting items for systematic reviews and meta-analyses guidelines (PRISMA).

### Inclusion criteria

We will include studies that assess:

1. Participants: patients with breast cancer receiving medical treatment.
2. Intervention: omega-3 fatty acids supplementation
3. Comparison: Any control group or other treatment.
4. Types of studies: randomized controlled trials with no publication date restriction

### Exclusion criteria

1. Full-text documents not available
2. The following study designs: cross-sectional studies, case control, cohort, case reports, case series, letters to the editor, editorial, narrative review, systematic reviews, correspondence, short communications, technical notes, commentaries and pictorial essays.
3. Studies published in any language other than Spanish or English

### Literature search, data collection and coding

We will search the following databases: 1) PubMed, 2) Central Cochrane Library, and 3) Embase. Duplicates articles will be removed using Endnote software. Two reviewers will screen titles and abstracts, and will identify potentially relevant studies inclusion and exclusion criteria. Disagreements will be discussed with another reviewer. Then, we will screen full-text in a similar fashion.

The complete search strategy for each database, the number of hits retrieved, and the reasons for exclusion of any studies at the full text stage of selection will be recorded and provided as appendices. We will record the results of included and excluded publications and show them on a PRISMA flowchart^9^.

The data extracted from the selected articles will include:

- Study details: first author, corresponding author, article title, country, year of publication and year of data collection, duration of follow-up, disease outcome, dietary assessment, medical treatment.
- Subjects: number of participants, range of age, population source, location, inclusion and exclusion criteria of studies selected Study.
- Results: nutritional and clinical outcomes

In case we find a study in which the methodology or outcome is not clearly specified, we will try to contact the corresponding author. If we do not receive an answer, then the study will be excluded from the systematic review.

## Risk of bias (quality) assessment

We will assess risk of bias using the Cochrane Risk of Bias Tool for RCT and present a table with the results of this assessment^10^.

## Statistical analysis

The results of each study will be expressed, when possible, as standardized or weighted mean difference or relative risk (RR) with corresponding 95% confidence intervals for continuous or dichotomous data, respectively.

The included studies will be grouped into sub-groups of similar population, intervention, and outcome. If a subgroup of studies appears comparable, we will investigate the possibility of pooling such data via formal meta-analysis analytical techniques.

We will assess heterogeneity using an I² statistical. In the case of finding heterogeneity, we will use a random-effects model. The data will be processed with the software Stata v14.0.

## Data Availability

We will search the following databases: 1) PubMed, 2) Central Cochrane Library, and 3) Embase. The search terms will be adapted to each database.We will review the references of included studies in order to find more potential eligible studies.

## Conflicts of interest

All authors declare to have no interest conflict.

## Appendix

### Search strategy

#### Pubmed

(“Fatty Acids, Omega-3”[Mesh] OR “Omega-3”[tiab] OR PUFA[tiab] OR “Eicosapentaenoic Acid”[Mesh] OR Eicosapentaenoic[tiab] OR “Docosahexaenoic Acids“[Mesh] OR Docosahexaenoic[tiab] OR “Fish Oils”[Mesh] OR “fish oils”[tiab] OR DHA[tiab] OR EPA[tiab]) AND (“Breast Neoplasms”[Mesh] OR “Breast neoplasm”[tiab] OR “Breast cancer”[tiab])

#### Filter

randomized controlled trials

#### Embase

((‘omega 3 fatty acid’/exp OR ‘omega 3 fatty acid’ OR ‘fish’/exp OR fish) AND (‘oils’/exp OR oils) OR ‘dha’/exp OR dha OR epa OR pufa OR ‘omega 3’/exp OR ‘omega 3’ OR eicosapentaenoic OR docosahexaenoic) AND (‘breast cancer’/exp OR ‘breast cancer’ OR ‘breast neoplasm’)

#### Filter

publication type:article

### CENTRAL

(“Fatty Acids, Omega-3“[Mesh] OR “Fish Oils“[Mesh] OR Omega-3 OR PUFA OR Eicosapentaenois OR Docasahexaenoic OR fish oils OR DHA OR EPA) AND (Breast neoplast OR breast cancer OR Breast Neoplasms[Mesh])

## Notes

### Competing Interest Statement

The authors have declared no competing interest.

### Funding Statement

This study did not receive external funding

### Author Declarations

This is a systematic review protocol and no human subjects were involved in this project. Therefore, this study was classified as non-human subject research and no approval was needed from an IRB/ethics committee

